# People Behavior Changes in China during COVID-19 Pandemic

**DOI:** 10.1101/2020.05.15.20097667

**Authors:** Yongzhong Wu, Mianmian Huang, Guie Xie, Xiangying Chen

**Affiliations:** Department of Industrial Engineering, South China University of Technology, Guangzhou, China; KingMed School of Laboratory Medicine, Guangzhou Medical University, Guangzhou, China

**Keywords:** COVID-19, Containment, virus transmission, behavior changes

## Abstract

While most countries implemented measures to reduce people activities and gathering to contain the coronavirus, the outcome may vary significantly depending on how and to what extent people behaviors have been changed. We conducted a survey of 1,048 people in five major cities in China to track and quantify the behavior changes in different periods since the outbreak. It is found that there was nearly 80% reduction of out-of-home activities (working, eating, shopping, taking public transportation, and travelling) during the peak period. Such activities are gradually increasing after the easing of containment measures but still significantly below pre-outbreak level. The significant behavior changes have contributed to the rapid control of virus transmission in China. While countries are reopening the economies before the virus disappears, the system and capacity of testing and contact tracing should be carefully designed with the tracking of people behavior changes in the future.

## 1. Background

As of 10 May 2020, there has been more than 4 million confirmed COVID-19 cases worldwide since the first case reported in Wuhan, China in December 2019 [1]. The number of cases is still increasing rapidly, with more than 85,000 new cases on May 9. As the first country to report coronavirus cases, China has passed the peak of the first wave and there were only 15 new cases on May 9 [2].

Although it was reported that China has mobilized an unprecedented large-scale public health response [3], the exact impacts on people’s behavior changes were not known. Most strict measures were taken in the epicenter Wuhan and Hubei Province, including the suspension of outbound travel [4], while much less strict social isolation and screening measures were taken in most parts of the country (see Figure 1). In most part of China, people were not strictly forbidden from going out of home.

The cordon sanitaire of Wuhan and the control of cross-regional movement is shown to have helped mitigate the virus spread to other areas of China and the world to some extent [5, 6, 7]. Nonetheless, behavior changes and awareness of the disease in the population are most important to reduce disease transmissibility in the community [6]. While the cross-regional travel data have been conveniently retrieved from location-based services (LBS) on the internet (e.g., Baidu LBS) and used in the modelling the transmission process of COVID-19 [4, 6], the information and analysis on behavior changes of people in China is lacking.

**Figure 1:**
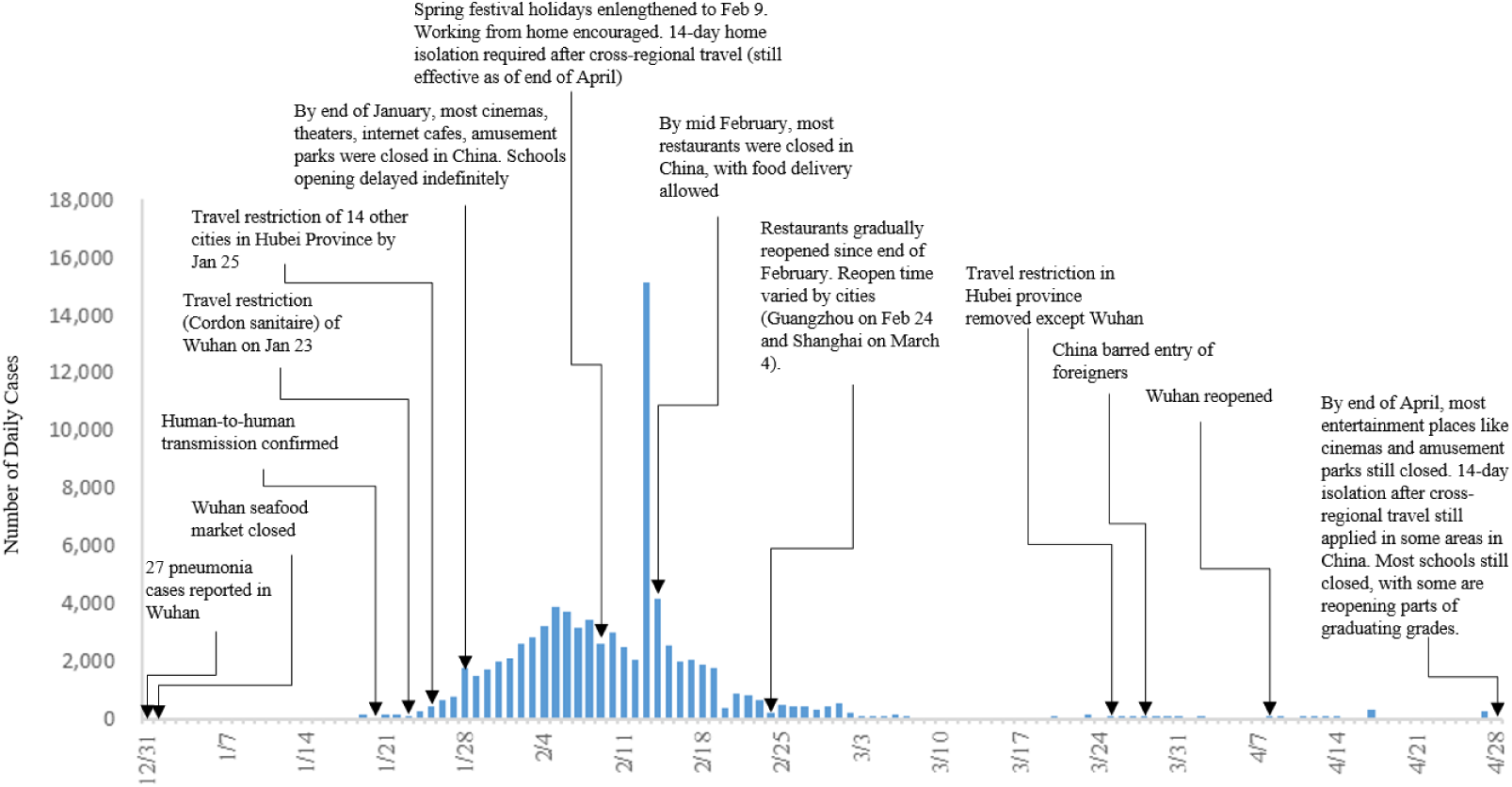
Timeline of Establishing and Easing of Containment Measures in China

## 2. Behavior Changes in China

We conducted a survey of 1,048 residents in major cities in China including Beijing, Shanghai, Guangzhou, Shenzhen, and Chongqing. The survey was conducted on a major survey platform (Tencent Questionnaire) during April 26-30, 2020. The respondents covered various occupancies, including white collar employees (41.0%), factory workers (11.9%), service industry employees (like shopping mall and restaurants) (10.1%), civil servants (2.1%), medical personnel (2.4%), self-employed (6.5%), freelancer (9.6%), students (8.9%), unemployed/retired (3.4%), and others (4.1%). The constitution of the respondents is considered to be well representative of the demographic characteristics in China.

The survey was designed to investigate people’s behavior changes in out-of-home activities, including working, eating, shopping, taking public transportation, travelling and mask wearing in different periods, i.e., before the outbreak, during the peak period (early February), and most recently (end of April). In the survey, the peak period was referred to the week of February 1016 because Chines New Year festival was officially postponed to February 9 and daily new cases were peaking during this week. Tracking behavior changes during this period is considered to be much representative of people response without the impacts of New Year holidays.

### Out-of-home working

Most of people stopped working during the peak period. Based on the survey results, only 26% of people (excluding students, unemployed and retired) were working outside home in early February (after the Chinese New Year holidays). 21% of people said they were working from home. Figure 2 shows the rate of out-of-home working for people of different occupations. It can be seen that during the peak period, the working rate of medical personnel was the highest (60.0%), followed by civil servant (36.4%) and service industry employee (35.8%).

By the end of April, 78% of people have returned to work (out of home). 22% of people either have not resumed working or they are working from home.

**Figure 2:**
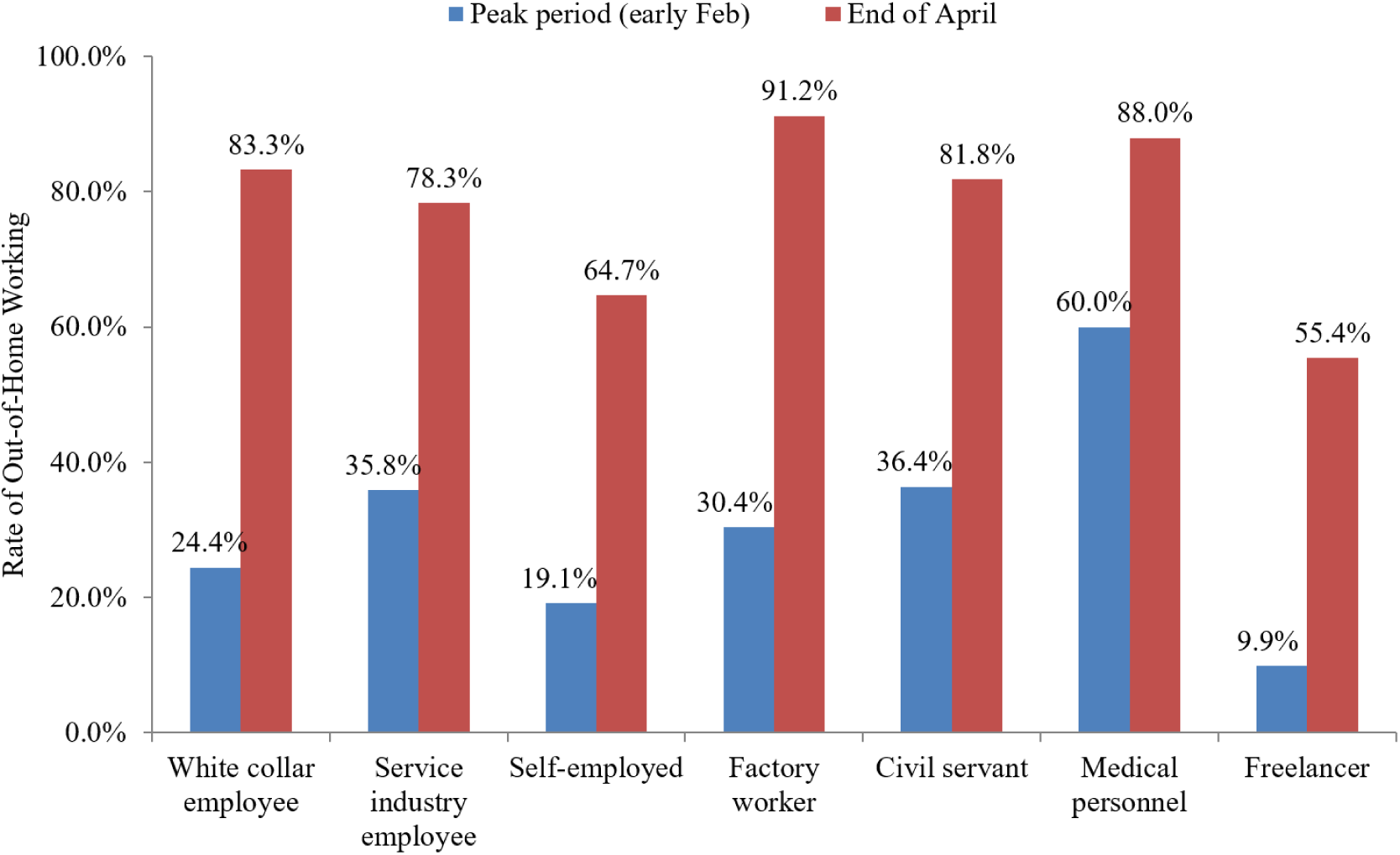
The rate of out-of-home working for people of different occupations

Besides working, common out-of-home activities which are associated to virus transmission have also been reduced significantly. Table 1 summarizes the changes of out-of-home eating and shopping, taking transportation, and traveling in different periods.

**Table 1:** Changes of out-of-home activities in China in different periods

|  |  | Percentage of people in different frequencies |  |  |  |  | Average Frequency (times per week) |
| --- | --- | --- | --- | --- | --- | --- | --- |
|  |  | 0 times per week | 1-2 times per week | 3-5 times per week | 6-10 times per week | >10 times per week |  |
| **Out-of-home Eating** | Before outbreak | 39% | 27% | 20% | 8% | 6% | 2.6 |
|  | Peak (Early Feb) | 90% | 6% | 2% | 1% | 1% | 0.4 |
|  | Late April | 52% | 29% | 10% | 5% | 4% | 1.7 |
| **Shopping** | Before outbreak | 18% | 44% | 30% | 6% | 3% | 2.6 |
|  | Peak (Early Feb) | 69% | 25% | 5% | 1% | 0% | 0.7 |
|  | Late April | 29% | 46% | 18% | 5% | 2% | 2.0 |
| **Public transportation** | Before outbreak | 41% | 15% | 14% | 11% | 19% | 3.9 |
|  | Peak (Early Feb) | 87% | 7% | 4% | 1% | 2% | 0.5 |
|  | Late April | 51% | 18% | 11% | 9% | 10% | 2.6 |
|  |  | 0 times per month | 1-2 times per month | 3-4 times per month | 5-6 times per month | >7 times per month | Average (times per month) |
| **Travelling** | Before outbreak | 64% | 30% | 4% | 1% | 1% | 0.6 |
|  | Peak (February) | 94% | 4% | 2% | 0% | 0% | 0.1 |
|  | April | 85% | 11% | 2% | 1% | 0% | 0.3 |

### Out-of-home eating and shopping

Before the outbreak, on average people went out eating (in restaurants) 2.6 times per week. This number significantly dropped to 0.4 time per week (85% reduction) during the peak in early February. 90% of people did not go out eating at all. At the moment (late April), people eat out more frequently than in the peak period. However, the frequency of eating still does not recover to the pre-outbreak level. The average frequency is 1.7 times per week now, which is 35% lower than the pre-outbreak level.

For out-of-home shopping (including shopping malls, stores and wet markets, etc.), the average frequency was 0.7 times per week during the peak, or 73% reduction comparing to 2.6 times per week before the outbreak. 69% of people did not go out shopping at all during the peak period, as most living supplies could be delivered via online shopping. Currently, people go out shopping 2.0 times per week, still 23% lower than the pre-outbreak level.

### Usage of public transportation

Usage of public transportation (bus and subway) has also been affected dramatically. Before the outbreak, as much as 59% of people were using public transportation (bus and subway) at least once per week. This number plummeted to only 13% during the peak in early February. Currently in late April, 49% of people are using public transportation.

On average, people (the whole population) used public transportation 3.9 times per week before the outbreak, but only 0.5 times per week (87% reduction) during the peak and 2.6 times per week at the end of April (33% reduction).

### Travelling

Due to the strict measures of cross-regional transportation, travelling has been dramatically minimized during the outbreak. In the survey, we asked the respondents about the monthly frequency of cross-city travelling for both leisure and working before the outbreak, in February, and in April. Before the outbreak, 36% of people travelled at least once per month. This number fell to 6% in February and 15% in April.

In terms of travelling frequency, People travelled 0.6 times per month before the crisis. The number significantly dropped to 0.1 time (83% reduction) in February, and recovered to 0.3 time (still 50% reduction) in April.

### Wearing mask

Although there has been a worldwide discussion on the universal use of face masks in the community [8], China government strongly promoted the public use of face masks to reduce virus transmission.

During the peak period, people used a mask in 92% of time in street, 79% of time in working place, and 95% of time in crowded environment (including shopping mall, wet market and public transport). It can be seen that the mask usage level was very high in both streets and crowded environment. The relatively low usage level in working place may be due to the shortage of masks in that period (21% of the respondents said that they once had no mask to wear at a certain time).

Currently, the mask usage still remains at a very high level. In late April, people used a mask in 90% of time in street, 83% of time in working place, and 93% of time in crowded environment. Figure 3 shows the photos of out-of-home activities in different public places in China. It shows that there is still a relatively high level of awareness and compliance to the containment measures in the public, including wearing masks and temperature testing.

**Figure 3:**
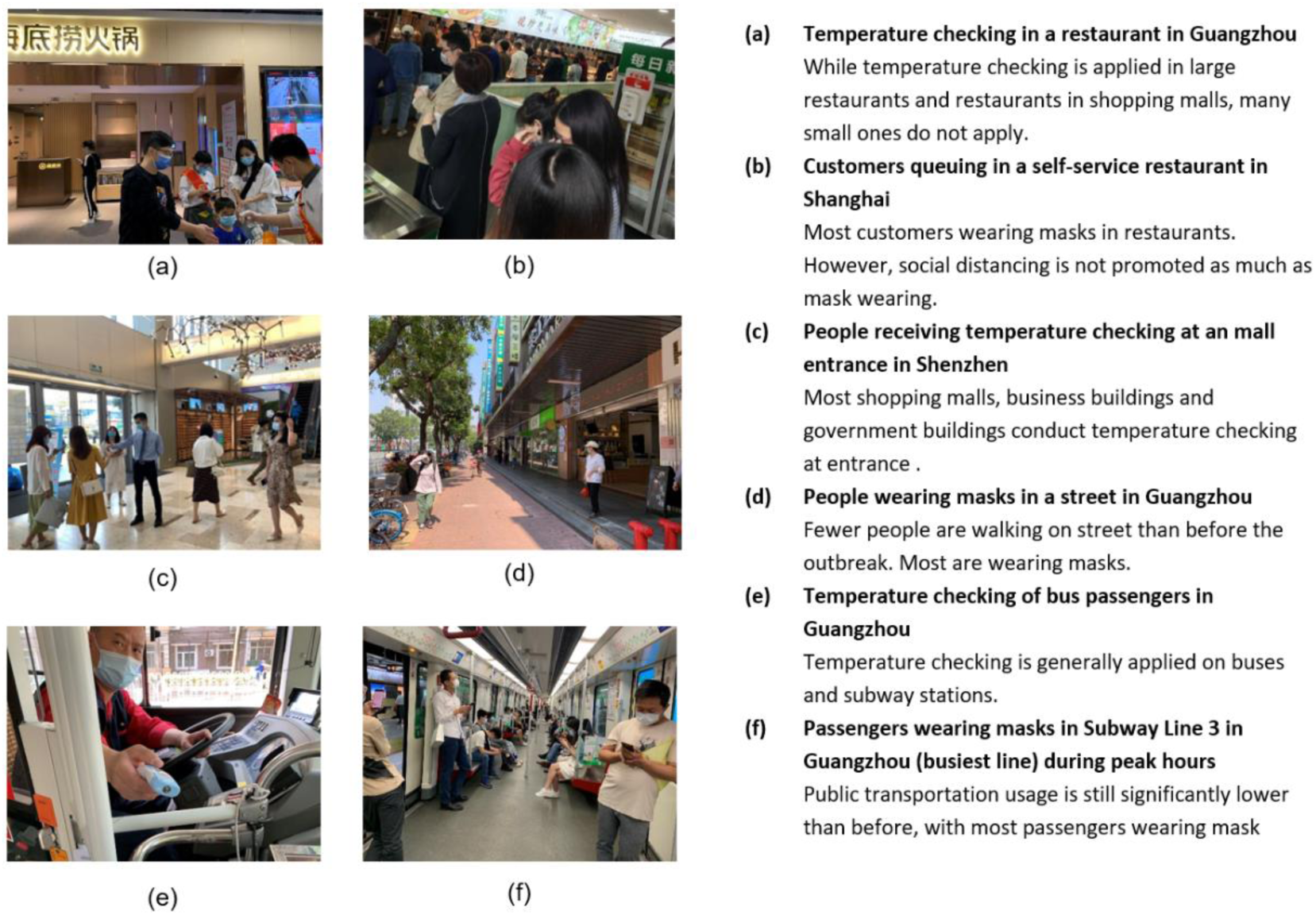
Photos of out-of-home activities in different public places in China

## 3. Discussion

Figure 4 summarizes the overall impacts of COVID-19 on different activities. During the peak period in early February, the overall out-of-home activity level was reduced to about 20% of the pre-outbreak level (average impacts of different activities). Since the reopening, the overall activity level gradually recovered to 67% of pre-outbreak level in late April. It is believed that the dramatic changes of people behavior contributed to the rapid containment of virus transmission in China.

**Figure 4:**
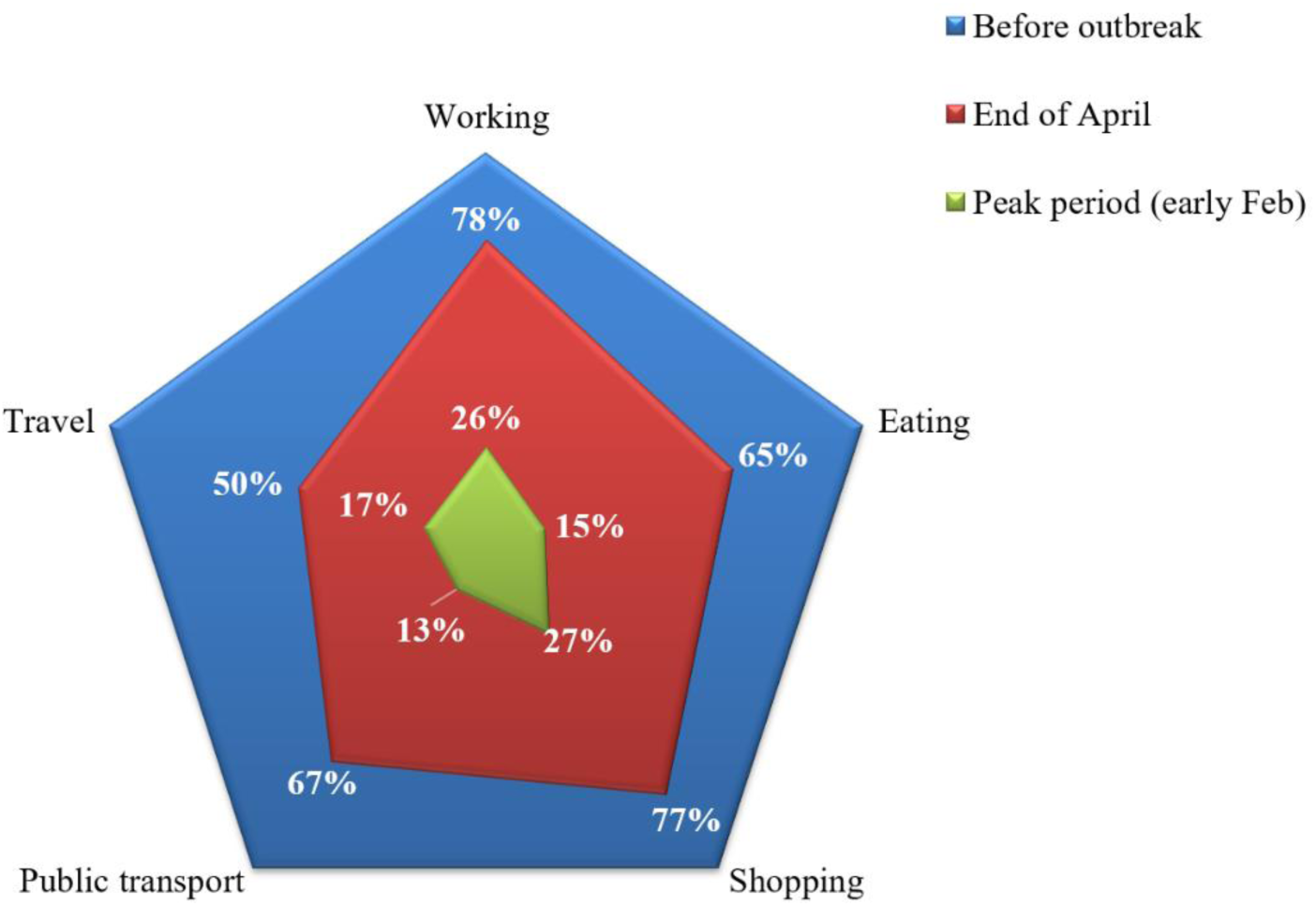
Level of behavior changes in different activities during the COVID-19 pandemic (in terms of frequencies)

Public transportation and out-of-home eating were affected most in the peak period (at 13% and 15% of out-of-break level). Working and shopping were least affected in the peak period (at 26% and 27% of pre-outbreak level) and recovered most rapidly after the peak (78% and 77% level). This may be due to the reason that working and shopping are essential to household living despite the risk of transmission. Travelling was also severely affected but recovered most slowly. Currently, the travelling frequency is still half of the pre-outbreak level as people try to avoid non-necessary travelling.

As people’s activities are still significantly lower than pre-crisis level and continue to recover in the coming future, there is a concern about a second wave of spread in China. Currently, Chinese government is much relying on wide testing and contact tracing to prevent a second wave. For instance, a Starbucks employee in Guangzhou was diagnosed with COVID-19 on April 18 and more than 2,300 people in potential contacts were tested [9]. This instance further triggered a great testing effort in the whole city. During the same weekend (April 18-19), Guangzhou Municipal Commission reported to have conducted more than 68,000 testings and identified 7 infection cases who were all asymptomatic [10]. While Guangzhou is one of the cities with greatest testing capacity, the risk of a second wave may vary in different cities in China.

Outside China, most countries are still implementing measures to reduce people out-of-home activities and gathering, the effectiveness of such measures may differ significantly depending on the compliance of people to such measures and the magnitude of behavior changes. The behavior changes of people should be factored in the modeling of the virus transmission process for both research and policy making. While countries are reopening the economies before the virus completely disappears, the system and capacity of the testing and contact tracing should be carefully designed with the tracking of people behavior changes in the future.

## Data Availability

all data referred to in the manuscript are available.

## Author Contributions

Wu YZ conceptualized the original idea of this paper, designed the survey questionnaire, led the main analysis, wrote the first draft of the paper and reviewed the final version. Huang MM conducted the analysis of the survey results. Chen XY implemented the survey on the survey platform and collected the results. Xie G refined the analysis and discussion in the paper.

## Statement of Ethical Approval

Ethical approval is given by South China University of Technology.

